# Efficacy of *Wolbachia*-mediated sterility for control of dengue: emulation of a cluster randomized target trial

**DOI:** 10.1101/2023.11.29.23299172

**Authors:** Jue Tao Lim, Diyar Mailepessov, Chee Seng Chong, Borame Dickens, Yee Ling Lai, Youming Ng, Lu Deng, Caleb Lee, Li Yun Tan, Grace Chain, Soon Hoe Ho, Chia-Chen Chang, Pei Ma, Somya Bansal, Vernon Lee, Shuzhen Sim, Cheong Huat Tan, Lee Ching Ng

## Abstract

**Background:** Matings between male *Aedes aegypti* mosquitoes infected with *w*AlbB strain of *Wolbachia* and wildtype females yield non-viable eggs. We evaluated the efficacy of releasing *w*AlbB-infected *Ae. aegypti* male mosquitoes to suppress dengue.

**Methods:** We specified the protocol of a two-arm cluster-randomised test-negative controlled trial (cRCT) and emulated it using a nationally representative test-negative/positive database of individuals reporting for febrile illness to any public hospital, general practitioner or polyclinic. We built a cohort of individuals who reside in *Wolbachia* locations versus a comparator control group who do not reside in *Wolbachia* locations. We emulated a constrained randomisation protocol used in cRCTs to balance dengue risk between intervention and control arms in the pre-intervention period. We used the inverse-probability weighting approach to further balance the intervention and control groups using a battery of algorithmically selected sociodemographic, environmental and anthropogenic variables. Intention-to-treat analyses was conducted to estimate the risk reduction of dengue given *Wolbachia* exposure.

**Findings:** The final cohort consisted of 7,049 individuals residing in areas treated by *Wolbachia* interventions for at least 3 months and 69,216 individuals residing in non-treated areas in the same time period. Intention-to-treat analyses revealed that, compared with controls, *Wolbachia* releases for 3, 6, 12 or more months was associated to a 47% (Odds ratio (OR): 0.53 [0.45-0.62]), 47% (OR: 0.53 [0.50-0.65]) and 59% (OR: 0.41 [0.39-0.50]) protective efficacy against dengue respectively. When exposed to 12 or more months of *Wolbachia* releases, protective efficacy ranged from 36% (OR: 0.64 [0.58-0.96]) to 77% (OR: 0.23 [0.22-0.33]) dependent on township, and from 48% (OR: 0.52 [0.48-0.7]) to 78% (OR: 0.22 [0.09-0.32]) across years. The proportion of virologically confirmed dengue cases was lower overall in the intervention arm, and across each subgroup. Protective efficacies were found across all townships, years, age and sex subgroups, with higher durations of *Wolbachia* exposure similarly associated to greater risk reductions of dengue.

**Interpretation:** Our results demonstrated the potential of *Wolbachia*-mediated sterility for strengthening dengue control in tropical cities, where dengue burden is the greatest.

**Funding:** This study was supported by funding from Singapore’s Ministry of Finance, Ministry of Sustainability and the Environment, National Environment Agency, and National Robotics Program. JTL is supported by the Ministry of Education (MOE), Singapore Start-up Grant. SB is supported by an MOE Tier 2 grant.

## Introduction

Dengue is the most widespread arboviral disease worldwide and has shown sustained increases in burden year on year. The Americas and Southeast Asia routinely account for the majority of global cases^1^. Vector control remains the primary tool for mitigating the spread of dengue due to the lack of available therapeutics and highly effective vaccines globally. Conventional vector control measures include environmental management, source reduction and insecticide use^2,3^. While these measures can reduce the burden of dengue, they are resource intensive and yield diminishing returns.

*Aedes aegypti* is the primary vector for dengue. Yet, few randomized controlled trials have been conducted for control of vector populations or vector competence to reduce dengue transmission. Only one trial has used the endpoint of virologically confirmed dengue to examine the impact of introgressing “virus-blocking” strains of *Wolbachia* into field populations of *Ae. aegypti* on dengue incidence in Yogyakarta^4^.

A separate approach employs the use of incompatible insect technique (IIT), which encompasses release of only *Wolbachia*-infected male mosquitoes. Due to cytoplasmic incompatibility^5,6^, matings between *Wolbachia*-infected males and uninfected females yield non-viable eggs. Repeated releases of *Wolbachia*-infected males are thus expected to suppress wildtype mosquito populations and reduce disease transmission. IIT avoids disadvantages associated with traditional vector control, including genetic or behavioural resistance to insecticides, off-target effects, and the inability to locate cryptic larval sites. IIT further avoids fitness costs arising from exposure to male-sterilizing irradiation, which can reduce mating competitiveness of sterile males in a full sterile insect technique (SIT) program^7^. However, imperfect sex-sorting may lead to stable establishment of the released *Wolbachia* strain in the field due to unintentional release of fertile *Wolbachia*-infected female mosquitoes^8^. While this confers a reduced ability for the *Wolbachia*-established population to transmit dengue (a phenomenon exploited by the Yogyakarta trial above^4^), introgression renders cytoplasmic incompatibility-based IIT ineffective^8^.

To augment vector control in Singapore, we have conducted extensive field trials of *Wolbachia-*mediated IIT targeting *Aedes aegypti*. To reduce the likelihood of stable establishment, we combined IIT with SIT using low-dose irradiation to sterilize residual females during releases of *Wolbachia*-infected males^9^. As data from randomized trials are not available, observational analyses may be used to ascertain intervention efficacies by adopting a target trial emulation approach.^12-14^ This study used a nationally representative test-positive/negative cohort comprising individuals who were tested for dengue via all public hospitals, polyclinics and general practitioners to emulate a cluster-randomized test-negative target trial to ascertain the intervention efficacy of *Wolbachia-*mediated sterility to reduce the incidence of virologically confirmed dengue in Singapore.

## Methods

### Specification of the cluster-randomized test-negative target trial

We specified a cluster-randomized test-negative target trial^**10**^ to evaluate the efficacy of releasing *w*AlbB-infected *Ae. aegypti* male mosquitoes for dengue control via vector population suppression, from epidemiological week (EW) 1 2019 – EW 26 2022 in Singapore. The target trial was emulated using test-positive/negative databases, which comprised all patients who report to any general practitioner clinic, polyclinic or public/private hospital and were suspect of dengue illness during the trial duration in Singapore.

### Emulating randomisation protocols from cluster-randomised trials for *Wolbachia* interventions

26 townships in Singapore which were not subject to *Wolbachia* interventions were considered potential locations as control sites. Towns were demarcated planning areas used by government ministries and departments for administrative purposes. While four long-term *Wolbachia* field trial townships were not randomly pre-selected, we emulated constrained randomisation protocols for cluster-randomised trials by randomly selecting a set of 12 control townships, such that the historical dengue incidence of the intervention arm is similar to that of the control arm in the pre-intervention period^4,11^.This further prevents chance-imbalance in baseline dengue risk due to the small number of intervention (n=4) locations considered (See Supplementary Information). All locations practiced the same baseline dengue control protocol in the pre- and post-intervention periods^2,3^.

### Cohort

Under the Infectious Diseases Act, all laboratory-confirmed cases of dengue are legally mandated for reporting in the national dengue surveillance system. Approval from the Director General of Health, Ministry of Health, was obtained to collect and use the data of dengue-suspected patients, whose blood samples are sent for dengue tests, through a national network of diagnostic laboratories that support private clinics, public polyclinics, or public/private hospitals.

This project was exempted from formal bioethics review as it is not considered human biological research, as advised by the Ministry of Health, Singapore. All laboratory tests were performed for clinically directed reasons, and the data from these tests is routinely collected as part of routine dengue surveillance under the Infectious Disease Act, which exempts the need for informed consent.

In Singapore, 133,821 individuals reported for febrile illness and were tested for dengue at the Environmental Health Institute, hospital laboratories and commercial diagnostic laboratories, through general practitioner clinic, polyclinic or public/private hospital from EW1 2019 – EW 26 2022. All dengue-suspect patients were tested using either using an internally controlled RT-qPCR assay, dengue non-structural protein 1 (NS1) or IgM as diagnostic assays to detect dengue virus in serum samples^3,11^. We excluded individuals who were tested on more than one occasion in 4 weeks, individuals who had more than one residential address in different control or intervention townships and individuals who had been tested at different labs with conflicting dengue results. We also excluded individuals who had residential addresses at intervention sites at the time of the test but had not been exposed to *Wolbachia* interventions for at least 3 months, based on exposure criteria described below.

Four intervention townships (Bukit Batok, Choa Chu Kang, Tampines, Yishun) had 7,049 tested individuals included in the study period. After constrained randomisation, we selected 12 control townships (Bedok, Bishan, Clementi, Geylang, Jurong West, Kallang, Pasir Ris, Punggol, Queenstown, Sengkang, Toa Payoh, Woodlands) with 69,216 tested individuals in the study period. The control arm had an average dengue incidence rate normalized by population size which was less than 5% different from the intervention arm in the pre-intervention period of EW1 2010 to EW52 2016, indicating good balance in historical dengue risk between arms.

### Outcomes of interest

Analysis considered *Wolbachia* exposure as a binary classification based on the home address of the individual in an intervention sector within an intervention township (*Wolbachia*-exposed) or a control sector within the selected control townships (*Wolbachia*-unexposed). Sectors comprise 10 or more public housing apartment blocks and measured around 0.088 km^2^ on average and are used for planning of surveillance and control for environmental infectious diseases in Singapore.

We subcategorised *Wolbachia* exposure based on whether an individual resides in a sector which experienced sustained *Wolbachia* releases for 3, 6 or 12 or more months, due to the time required for releases to induce noticeable vector suppression (See supplementary information). Home address was defined as the primary place of residence reported on the diagnostic test date. The intervention effect was estimated from an odds ratio comparing the exposure odds (residence in an intervention location for 3, 6 or 12 or more months) among participants who were dengue test-positive versus test-negative controls, with the use of logistic regression (See statistical analysis below). The null hypothesis was that the odds of residence in an intervention sector would be the same among participants who test-positive as that among test-negative controls. Intervention efficacy was calculated as 100×(1−odds ratio).

### Characterisation of intervention

Male *Wolbachia*-infected *Ae. aegypti* were released twice weekly (weekdays, 0630–1030 hrs) at four townships in high-rise public housing estates covering 607,872 individuals as of Epidemiological Week (EW) 26 2022. Bukit Batok, Choa Chu Kang and Yishun towns were subjected to interventions which combined IIT with SIT. Tampines town used the high-fidelity sex-sorting methodology and also progressively adopted SIT protocols to release irradiated mosquitoes from January 2020^9^. To trial whether *Aedes aegypti* population suppression could be sustained over increasingly larger areas, an expanding release strategy was adopted in two large towns (Yishun, Tampines), where release sites were gradually expanded to adjacent neighbourhoods. In Bukit Batok and Choa Chu Kang towns, a targeted release approach was adopted, which focused releases on areas with high *Aedes aegypti* abundance and persistent dengue transmission. (Table 1, See supplementary information for full details). Adult *Aedes aegypti* populations in release and control sites were monitored using Gravitraps, with an average of six Gravitraps deployed per apartment block^12^.

**Table 1:**
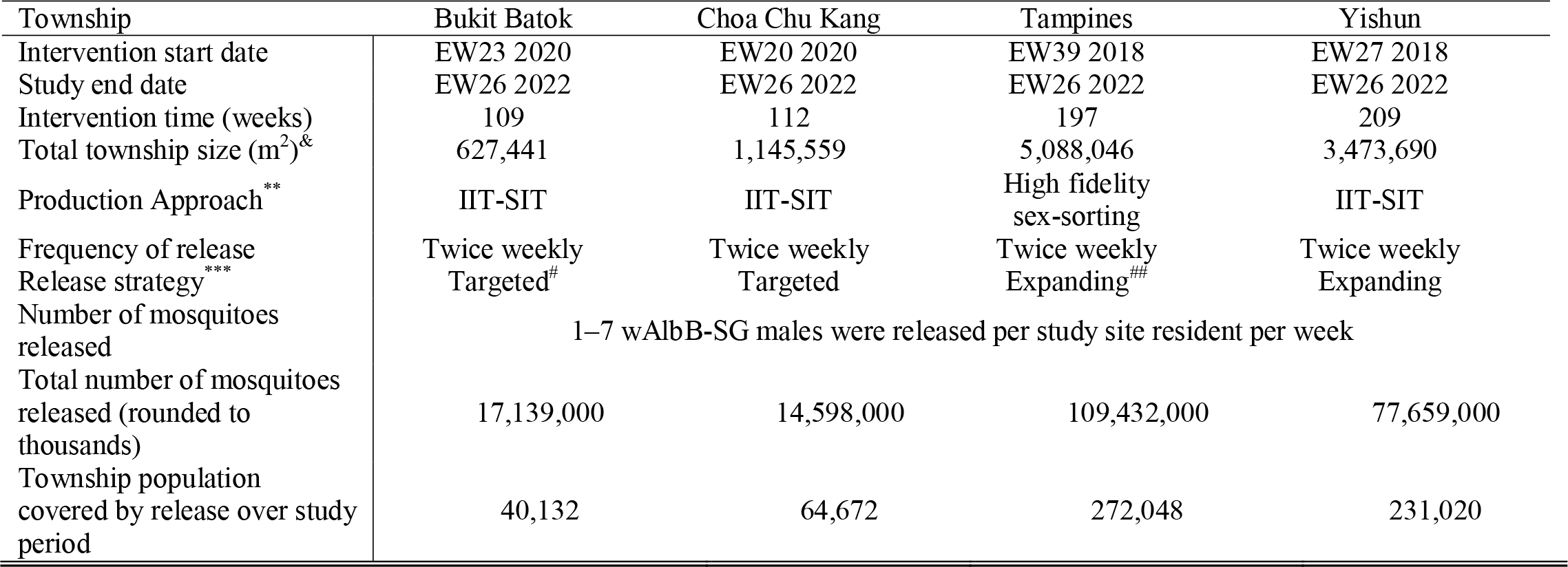
Summary of *Wolbachia* intervention approaches over 4 townships. ^&^total area of public housing estates subject to release in respective townships in EW26 2022 ^*^Computed as (sum of area of releases multiplied by weeks of release till end of study period) over (total area of township multiplied by total release weeks). Areas were considered covered once they receive at least 6 months of Wolbachia interventions. ^**^The IIT-SIT approach and high-fidelity sex-sorting were detailed in supplementary information section 1 and has been previously characterised^9,13^. ^***^denotes approach to releasing Wolbachia-infected males ^#^Targeted approach which focused releases on areas with high *Aedes aegypti* abundance and persistent dengue transmission ^##^Expanding (“rolling”) approach where release sites were gradually expanded to adjacent neighbourhoods

### Covariates

We extracted a comprehensive set of spatially explicit variables to characterize environmental heterogeneity across sectors. Covariates considered prior to variable selection include, (1) vegetation maps with areas classified across multiple vegetation types including forest and managed vegetation to signify availability of natural breeding sites and nectar availability for male mosquitoes. (2) The averaged Normalized Difference Vegetation Index per sector, as an alternative measure of vegetation (3) To represent host density and urban breeding habitat availability, data on the locations of all public housing estates where over 75% of Singapore’s resident population reside were obtained. Utilising residential location and resale data, the average age of public housing residences was collected as older age is a well-established risk factor for higher mosquito abundance^14^.(4) Average residence price over the study duration as a proxy for household income and socioeconomic status. (5) Building height was calculated according to the number of floors with an average height of 3m. (6) The number of condominiums/landed properties was collected within each sector representing additional hosts being available. The percentage cover of built area was calculated as a sum of all residential, commercial and industrial buildings, representing the level of urbanicity, which has been associated with *Aedes aegypti* presence^14^. (7) The major open drainage network for Singapore was obtained from the Public Utilities Board and has been previously shown as a key breeding site for mosquitoes around public housing apartments^15^. The average distance of each block within a sector to a drain was measured as well as the length of the network within the sector. (8) Well-established meteorological variables which are known to affect mosquito survival or fecundity were collected. These included daily mean, maximum, and minimum temperature, total rainfall, and wind speed, which were obtained from 21 local weather stations. Hourly dewpoint and ambient ground air temperature were also taken from remote sensing measurements to estimate relative humidity over the time period using standard formula. These values were aggregated at a weekly level to correspond with the temporal frequency of dengue test-positive/negative data. Data sources and processing procedures were explicitly detailed in the Supplementary Information.

### Statistical analysis

Baseline characteristics of the cohort were presented as mean and standard deviation or as frequency and percentage. Standardized mean differences (SMD) were used to evaluate balance between intervention and non-intervention arms, with SMD <0.1 indicating good balance.

To estimate the effect of *Wolbachia* interventions on the risk of dengue, we employed a doubly-robust logistic regression framework. First, to estimate propensity score models, we used the considered set of covariates described above as the independent variables and *Wolbachia* exposure for at least 3,6 and 12 or more months as separate outcomes of interest. Prespecified variables included age and gender, with other factors selected using high-dimensional regression and additional trimming of highly multicollinear covariates (See supplementary information). Sensitivity analyses later also indicated that variable selection procedures did not influence *Wolbachia* protective effect estimates.

We adjusted for differences in baseline characteristics and the propensity to be selected as a treatment site between intervention/non-intervention arms through inverse probability weighting, incorporating the selected covariates. A propensity score of belonging to the intervention arm was computed using a logistic regression that used the abovementioned covariates as explanatory terms. Inverse probability weights were computed as 1/propensity score for tested individuals who were *Wolbachia*-exposed; and 1/(1–propensity score) for tested individuals who were not *Wolbachia*-exposed. SMDs were used to assess covariate balance after inverse probability weighting. Thereafter, odds ratios (ORs) of being dengue test-positive between the intervention and control groups were estimated using a logistic regression model, with inverse probability weights applied. A doubly robust approach was employed for this model, where covariates used to construct inverse probability weights were included in each model specification as explanatory variables. This approach was used to prevent model misspecification in the generation of inverse probability weights or ORs in analyses.

To account for within-sector dependencies, we relied on cluster bootstrap based on 1,000 clustered resamples. Balanced bootstrap resampling based on sector membership can account for within-sector dependencies and has been used as a competitive approach to analyse hierarchical data^16^. The associated bootstrap percentile-based confidence interval was used to construct the 95% confidence interval for odds ratios, and findings were considered to be statistically significant when the 95% confidence intervals for ORs did not cross 1.

### Subgroup analysis

We repeated all analysis by subsetting to intervention townships and specific years (2019, 2020, 2021, 2022) to examine any potential differences in intervention effect by location, and between epidemic and inter-epidemic years. We also repeat analysis by age (<20, 20-65, 65+) and sex (male, female) subgroups as dengue risk may be mediated by immunity levels in each age group or gender. Here, we conducted subgroup analyses by re-estimating odds ratios using the aforementioned statistical procedures, but only using individuals within that specific subgroup.

### Robustness checks

We conducted a battery of sensitivity analyses to ensure the robustness of our model estimates. We (**1**) repeated all analysis without adjustment for covariates in the main logistic regression step (**2**) re-randomised our allocation of controls 1000 times and repeated our analysis by using the newly allocated controls arm, and compared our primary estimate of intervention efficacy against the empirical distribution of re-randomised intervention efficacies (**3**) used the full set of covariates, instead of the pre-selected covariates in our main analysis, to recompute odds ratios and intervention efficacies (**4**) we conducted in-space placebo checks on control sites, taking each allocated control site as the allocated placebo-intervention site and re-estimated odds ratios and intervention efficacies by comparing test-negative and positive individuals in the placebo-intervention versus other control sites (**5**) we conducted placebo checks on intervention sites, taking each intervention site in 52 and 104 weeks before the actual intervention as the intervention period, and recomputed intervention efficacies among allocated controls and interventions based on this placebo-intervention period. (**6**) We recomputed intervention efficacies using cluster bootstrap on the cluster rather than sector resolution.

## Results

### Suppression of *Aedes aegypti* populations in field trial sites

Suppression of adult wild-type *Ae. aegypti* populations was demonstrated across the four field trial sites, with the Gravitrap *Aedes aegypti* Index (GAI) reduced as *Wolbachia* coverage increased across each township. When >50% coverage was achieved by EW 1 2022, the town-level GAI plunged below 0.05 for all sites (Figure 1, Supplementary Information).

**Figure 1:**
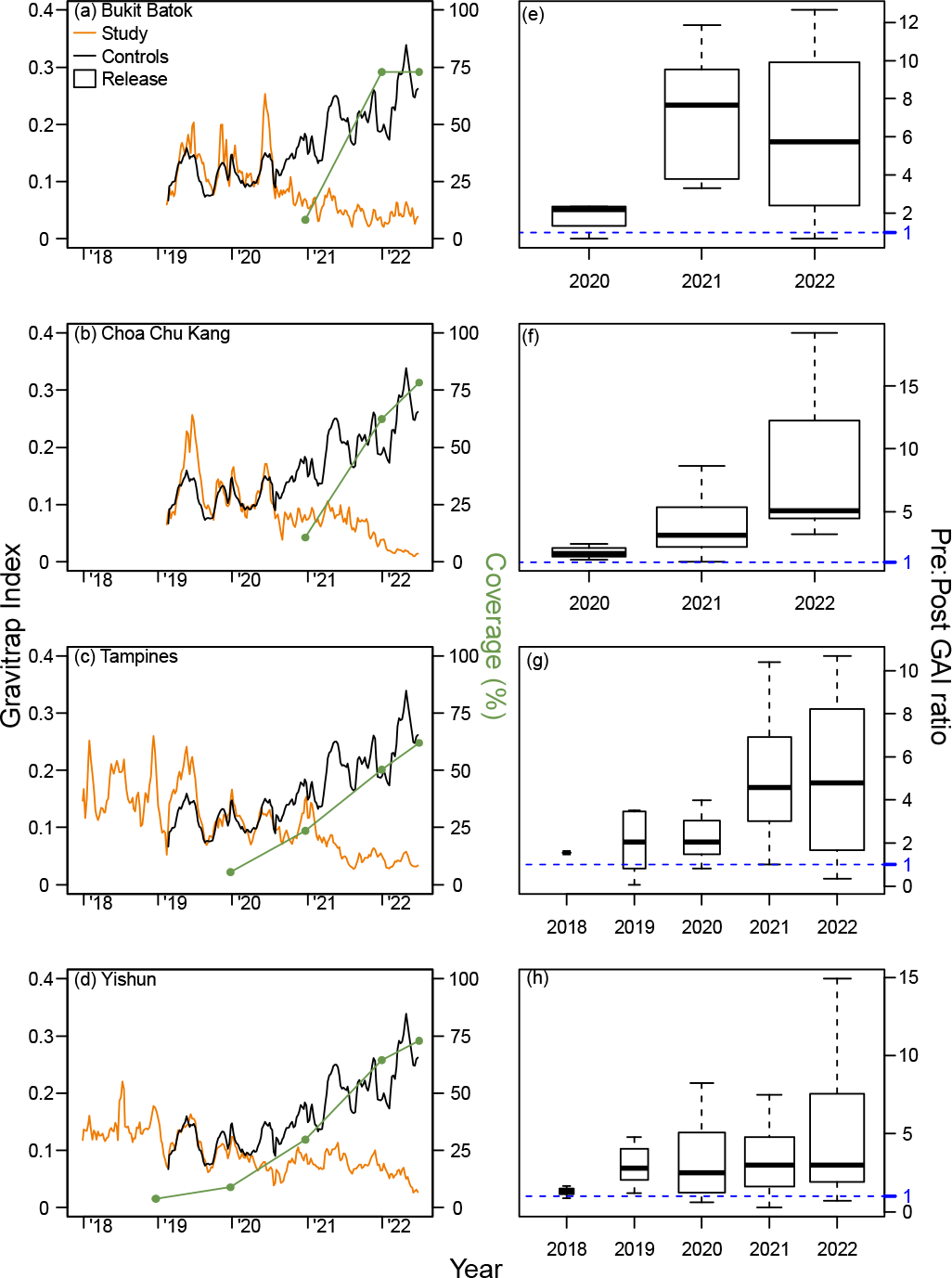
Weekly average, town level Gravitrap *Aedes aegypti* index (GAI) and *Wolbachia* coverage (%) from 2018 to 2022 in the intervention sites of (a) Bukit Batok, (b) Choa Chu Kang, (c) Tampines and (d) Yishun. The average GAI from the 12 controls is plotted for comparison. GAI is defined as the mean number of female adult *Ae. Aegypti* caught per functional Gravitrap per week, hence proxies for adult Ae. aegypti abundance in public housing areas in release area of each town. The geographical coverage (%) represents the percentage of areas within the town which is covered by *Wolbachia* interventions for at least six months and is calculated at the end of each year. Points represent coverage of *Wolbachia* interventions by the end of each year. The six-month mark for coverage is based on the time it takes *Wolbachia* release to have around 80% suppressive efficacy on *Ae. aegypti* abundance. The corresponding ratios of pre- and post GAIs at the sector is also plotted in (e), (f), (g) and (h), per year, on a per sector basis. The threshold of 1 indicates no difference between pre and post-GAIs for a specific sector in that specific year versus the pre-intervention period, and values above 1 indicate lower GAIs in the post-intervention period

### Study Characteristics

Among 133,821 individuals who reported for febrile illness from EW1 2019 – EW26 2022, in the intervention and control arms, 76,265 (56.99%) were included in the study. Baseline demographic and spatial-temporal characteristics between *Wolbachia*-exposed and unexposed groups before and after inverse probability weighting were presented in Table 2. Characteristics were well-matched after inverse probability weighting (Table 2) with small differences in baseline characteristics between both groups.

**Table 2:**
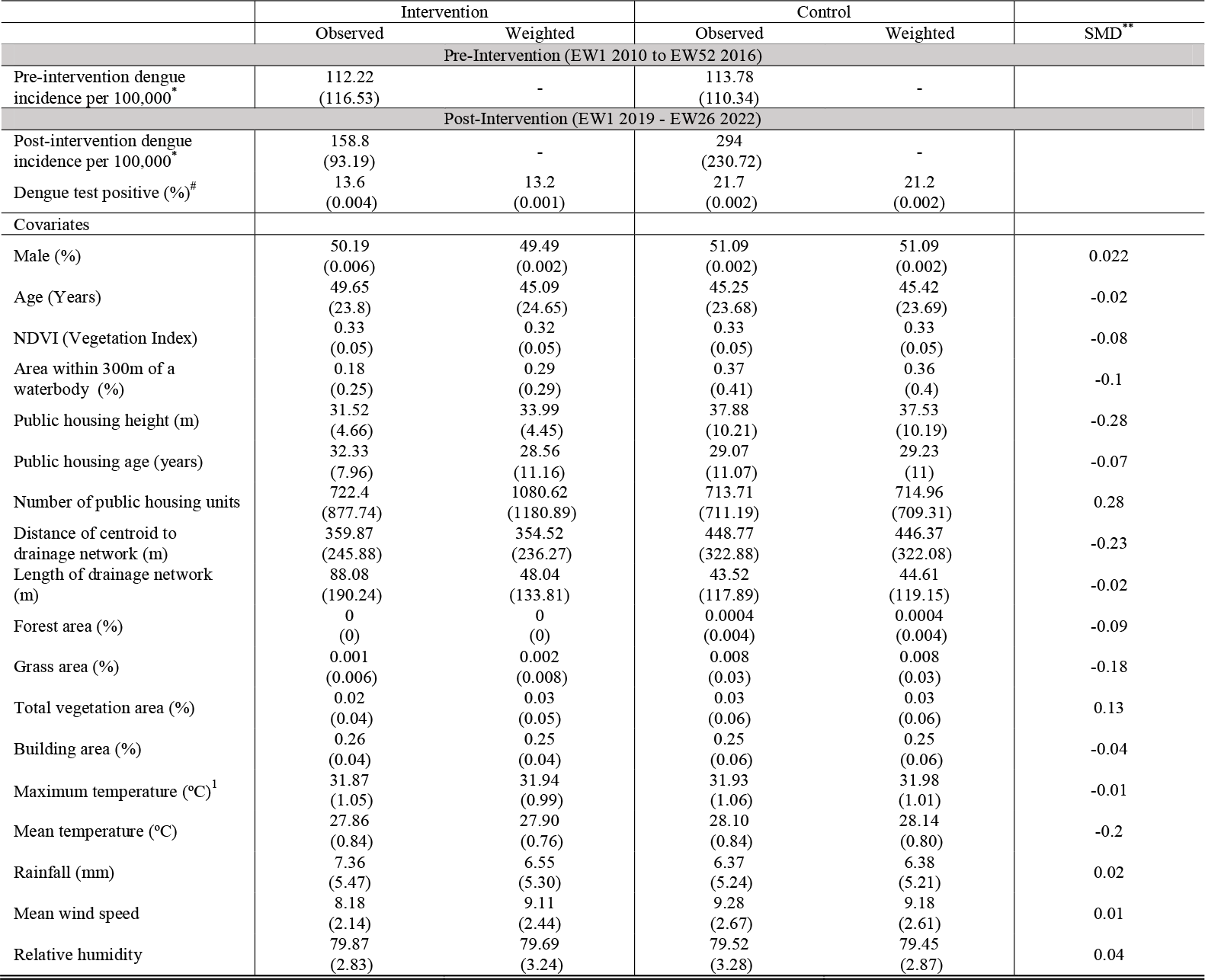
Baseline characteristics of study population pre- and post-*Wolbachia* releases in intervention and pre-selected control group, at the sector resolution. The numbers in bracket represent standard deviation for each characteristic. ^*^Pre-intervention period dengue incidence denotes number of dengue cases per 100,000 per sector annually ^#^Post-intervention percentage of dengue test positives compared to total number of tests per sector. Only data on dengue tests were available 2016 onwards. ^1^Maximum temperature was calculated by taking maximum of temperature across all sectors within intervention or control groups. Length of drainage network and number of public housing units were calculated by taking sum across all sectors within intervention or control groups. The remaining characteristics were calculated by averaging across all sectors within intervention or control groups. All the calculations were done for the specified time period. ^**^Standardized mean differences (SMD) after inverse probability weighting of intervention (*Wolbachia*-exposed) and controls (*Wolbachia*-unexposed) individuals. Tested individuals were considered *Wolbachia*-exposed here if they reside in a place of residence which has sustained Wolbachia interventions for 3 or more months.

### Efficacy of *Wolbachia*-releases in reducing risk of dengue

Among dengue-tested individuals residing in areas which were *Wolbachia-*exposed for more than 3 months, the percentage of individuals who tested positive for dengue (13.6%, 956 of 7,049 individuals) was lower compared to the *Wolbachia*-unexposed (21.7%, 14,986 of 69,216 individuals) group.

In primary analysis, *Wolbachia* exposure for 3, 6, 12 or more months was associated to a lower risk of being test positive for dengue. Higher periods of exposure associated to greater levels of protective efficacy – at 47% (Odds ratio (OR): 0.53 [0.45-0.62]), 47% (OR: 0.53 [0.50-0.65]) and 59% (OR: 0.41 [0.39-0.50]) for 3, 6 and 12 of more months of *Wolbachia* exposure respectively. Protective efficacies were higher in epidemic years (2019, 2020, 2022) versus inter-epidemic years (Table 3).

**Table 3:**
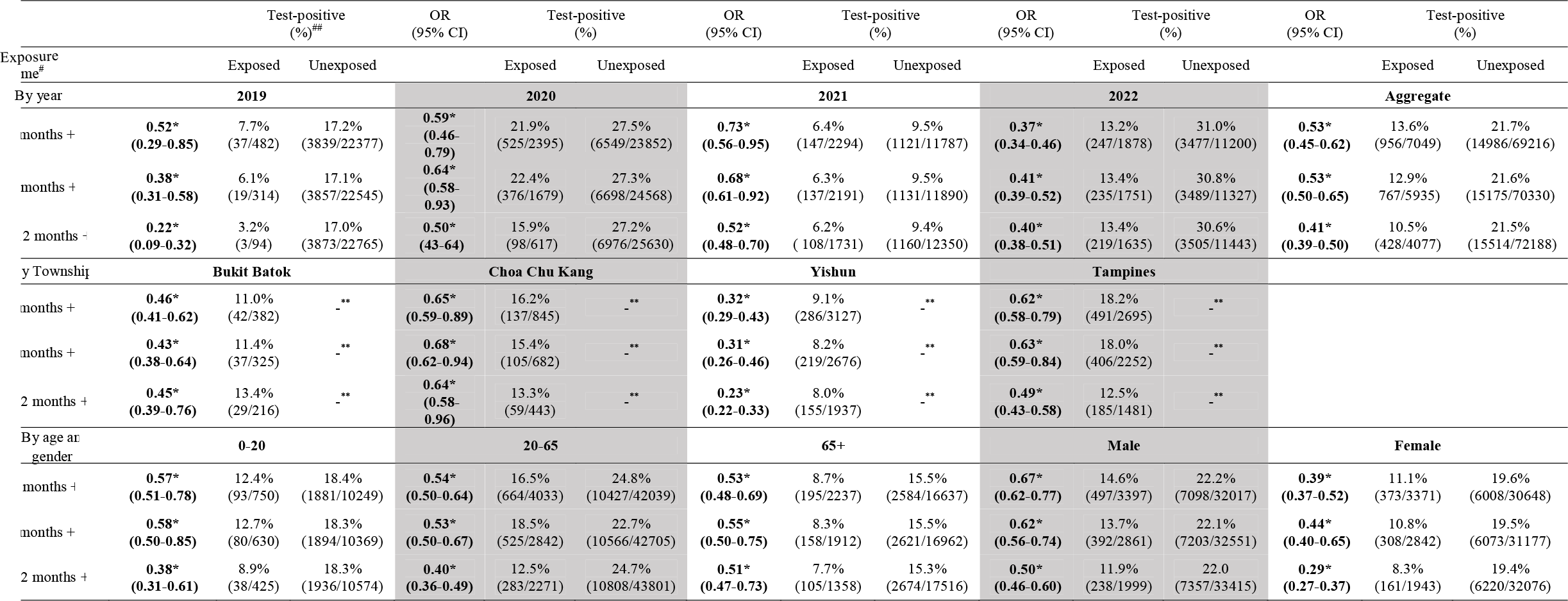
Odds ratios (OR) and test positive percentages for different *Wolbachia* exposure categories and across year, town, age and sex subgroups. ^*^Denotes an OR < 1 with 95% confidence intervals (CI) which are not bounded by 1 denotes a significant protective effect of *Wolbachia* interventions on the risk of dengue. ORs are estimated using doubly robust logistic regression with weights for each individual estimated using inverse probability weighting. Cluster bootstrap at the sector resolution was used to obtain CIs to account for sector-specific spatial clustering of data and the intervention. ^**^Unexposed group taken as the pre-randomised set of 12 controls ^#^An individual testing for febrile illness is considered *Wolbachia*-exposed if the individual resides in a sector with 3,6 or 12 or more months of sustained *Wolbachia* release ^##^Unweighted percentages of individuals testing positive in *Wolbachia*-exposed and *Wolbachia*-unexposed sectors

By stratifying analysis across townships, years, age and sex subgroups and taking *Wolbachia* exposure at 12 or more months as the reference, the highest level of protective efficacies were found in Yishun township, at 77% reduction (OR: 0.23 [0.22-0.33]), and lowest in Choa Chu Kang township, at 36% reduction (OR: 0.64 [0.58-0.96]) when compared across townships. The highest level of protective efficacy was in the adolescent (62% protective efficacy, OR: 0.38 [0.31-0.61]) and adult age groups (60% protective efficacy, OR: 0.40 (0.36-0.49)) when compared across age groups and in females (71% protective efficacy, OR: 0.29 [0.27-0.37]) when compared against both sexes. Protective efficacies were found across all subgroups, with higher durations of exposure similarly associated to greater risk reductions of dengue – supporting consistent biologic replication of the intervention (Table 3).

### Sensitivity analyses

We conducted a battery of sensitivity analyses to ensure the robustness of our model estimates. Repeating all analysis without adjustment for covariates, or inclusion of all available covariates in main logistic regression step did not change intervention efficacy estimates. Re-randomising our allocation of controls into the control arm 1000 times and repeating our analysis by using newly allocated controls arms did not change intervention efficacy estimates versus our primary estimate of intervention efficacy. We further did in-space placebo checks on control sites, taking each allocated control site as the allocated placebo-intervention site and re-estimated protective efficacies by comparing test-negative and positive individuals in the placebo-intervention versus other control sites. There were few significant/positive intervention efficacies demonstrated in the placebo-control sites with the grand mean of OR estimates centered closely to 1. Lastly, we conduct in-time placebo checks on intervention sites, where we took placebo-interventions in the actual intervention site before the intervention began and demonstrated that there were no significant/positive protective effects in the intervention sites in the placebo-intervention period. Re-running our cluster bootstrap inference on the town, rather than sector level, also demonstrated that we could recover the intervention efficacy point estimates, albeit with wider confidence intervals due to the coarser spatial resolution used for bootstrapping. Explicit results on all sensitivity analyses are provided in the SI.

## Discussion

Releases of *w*AlbB-infected *Ae. aegypti* male mosquitoes reduced the incidence of dengue by 59% in areas which received 12 or more months of sustained intervention (Table 3). Across the four field trial sites, protective efficacies were heterogenous, with protective efficacies reaching 77% in Yishun township. Across all intervention townships, interepidemic and epidemic years, age and gender subgroups, the proportion of individuals with testing positive for dengue was lower versus the control arm, demonstrating consistent biological replication of the intervention effect (Table 3).

Lower protective efficacy in some towns, such as Choa Chu Kang (36%) may be due to a sizeable portion of sporadic cases which do not form part of local dengue clusters, indicating potential importation from non-intervention sites rather than acquisition within the release site (See SI). Prior work in the same study setting also examined the effect of *Wolbachia* on clustered/sporadic dengue case burdens, and estimated consistently low or negative protective efficacies for sporadic cases^17^. This suggested that a proportion of cases may have acquired the disease elsewhere, thus biasing the estimated protective efficacies for these townships downwards.

Variation across years was likely due to different dengue incidence rates at control sites, which are influenced by the national situation. Outbreak years (2019, 2020, 2022) typically yield higher efficacy than non-outbreak years (2021). The reduced efficacy following 2019 was also likely due to the expansion to Choa Chu Kang, which had lower efficacy, in the later years.

We emulated a cluster-randomised test-negative controlled target trial in this study, which has been employed to study the epidemiological efficacy of field interventions, such as *Wolbachia*^4,11^. By employing a large and representative cohort of patients who have suspect dengue illness and were tested for dengue, across all major diagnostic laboratories through public hospitals, general practitioner clinics and polyclinics, we were able to emulate a cluster randomized controlled target trial and incorporate key study characteristics, such as (**1**) constrained randomisation, which enabled intervention and control arms to have balanced dengue risk in the pre-intervention period, (**2**) the use of a test-positive and test-negative comparator groups, to avoid selection bias at the point of testing and enable detection of virologically confirmed dengue cases and thereby (**3**) enabling casual identification of the protective efficacy of *Wolbachia* on dengue.

The protective efficacy of *w*AlbB-infected *Ae. aegypti* male mosquitoes releases are consistent with previous laboratory and entomological field observations. Release of incompatible *Ae. aegypti* male mosquitoes can drive profound suppression of wild-type *Ae. aegypti m*osquitoes^8,13,18,19^. While previous field trials^4^ have demonstrated the protective efficacy of *w*Mel introgression in reducing dengue incidence, no study has yet examined the effect of incompatible insect technique in reducing risk of dengue. Our study combined data from large-scale field trial of *w*AlbB-infected *Ae. aegypti* male mosquitoes releases and utilized a robust cluster-randomized trial emulation framework to demonstrate the protective efficacy of the technology on dengue.

The technology has several key advantages. (**1**) while we have only demonstrated the protective efficacy of the approach for dengue, as it is the only *Aedes*-borne disease in constant circulation in the study setting, this efficacy should be similar against other diseases transmitted by *Ae. aegypti*, such as Zika, chikungunya and yellow fever, as the strategy aims to suppress vector populations rather than block disease transmission under the *Wolbachia* introgression approach. (**2**) Baseline studies demonstrate the high public acceptance towards the intervention^20^, which involves releases of non-biting males only. (**3**) Lastly, while dengue virus may evolve resistance to *Wolbachia* under the *Wolbachia* introgression approach^21^, our approach suffers no drawbacks related to *Wolbachia*-associated selective pressure of viruses.

Release of *Wolbachia*-infected male *Ae. aegypti* is a novel method for the control of dengue. It should however not be viewed as a replacement for baseline vector control methods. The technology can complement conventional approaches, such as source reduction, community engagement in further reducing dengue transmission. In our experience, the protective efficacy of the intervention on dengue, as well as its entomological efficacy are likely to be maximized if it is used to complement and enhance conventional vector control measures, rather than replace them.

## Data Availability

All data produced in the present study are available upon reasonable request to the authors

