## Supplementary Information for "Efficacy of *Wolbachia*-mediated sterility for control of dengue: emulation of a cluster randomized target trial"

**Supplementary Information: Efficacy of *Wolbachia*-mediated sterility to reduce dengue incidence: a target trial emulation study**

We first pre-specified a hypothetical cluster randomized controlled trial and the target trial emulation of the pre-specified trial (SI Table 1)

| Component | Hypothetical cluster randomized trial | Target Trial Emulation |
| --- | --- | --- |
| Eligibility | Patients who report for undifferentiated febrile illness to any general practitioner clinic, polyclinic or public/private hospital during trial duration and have home addresses which are in trial locations between EW1 2019 – EW26 2022 | Same as hypothetical trial |
| Intervention strategies | Release of wAlB-SG infected male *Ae. aegypti* in home address of participants. An individual is considered directly intervened by *Wolbachia* if home address resides in an area which has sustained releases for more than:   (1) 3 months (2) 6 months (3) 12 months | Same as hypothetical trial |
| Intervention assignment | Townships at historically high-risk of dengue transmission pre-selected to either receive intervention or control randomly.  Townships undergo a constrained randomisation strategy for control and intervention to prevent chance imbalance and have same historical dengue incidence rates in the pre-intervention period. | Four long-term *Wolbachia* field trial townships not randomly pre-selected.  Control townships undergo a constrained randomisation same as the hypothetical trial to match the historical dengue incidence rates of the intervention township in the pre-intervention period. |
| Blinding | Intervention and participants unblinded to intervention | Same as hypothetical trial |
| Follow-up | N/A. Repeated tests were removed | Same as hypothetical trial |
| Primary outcome | Test-positive for dengue | Same as hypothetical trial |
| Causal contrast | Intention-to-treat effect | Observational analogue of the intention-to-treat effect |
| Analysis Plan | Intervention (exposure) variable of interest: wAlB-SG infected male *Ae. aegypti* at home address will be considered as a binary classification based on time-since-intervention (See intervention strategies).  Outcome variable: is test-positive status for dengue at point of testing.  Doubly robust logistic regression will be used to assess the intervention effect by estimating the aggregate odds ratio, which compares the exposure odds among test-positive cases versus test-negative controls. The null hypothesis is that the odds of residing in the intervention locations are the same among test-positive cases as test-negative controls.  All analyses will be adjusted for environmental and anthropogenic risk factors associated with dengue transmission risk, and the propensity to receive treatment based on the same risk factors. See detailed statistical analysis plan below. | Same as hypothetical trial |

SI Table 1: Summary of the protocol of a target trial estimating differences in dengue transmission risk for individuals residing in *Wolbachia* release sites versus those in control locations who are not.

**Data**

We extracted a set of spatially explicit variables to represent environmental heterogeneity across sectors.

**Vegetation data** A 10m vegetation map^1^ with areas classified across multiple vegetation types including grass, forest and managed vegetation based on Sentinel-2 satellite data, was utilised to signify the availability of natural breeding sites and nectar availability for mosquito males. The percentage cover of each vegetation type was calculated within each sector as mosquitoes often show preferential areas to breed and rest. Similarly, the averaged Landsat Normalized Difference Vegetation Index per sector was also utilised for this purpose^2^.

**Residential data** To represent both host density and urban breeding habitat availability, data on the locations of public housing estates named Housing Development Board (HDBs) where over 80% of Singapore’s resident population reside was obtained from Onemap^3^. Utilising the HDB location and HDB resale data, the average age of HDB buildings was collected as older age is a well-established risk factor for higher Gravitrap indices^4,5^ This is due to building deterioration providing additional breeding habitats in cracks and design features such as laundry poles which are no longer built due to the pooling of water within the supports. Average HDB house price from 2015–2022, a proxy for household income and socioeconomic status, was calculated based on an XGBoost model previously employed^6^. Building height, which has also been correlated to Gravitrap indices, was calculated according to the number of floors and average height of each level of 3m. The number of condominiums and landed properties was additionally collected within each sector representing additional hosts being available. The percentage cover of built area was calculated as a sum of all residential, commercial and industrial buildings, representing the level of urbanicity, which has been associated with *Ae. aegpyti* presence^7^. The major open drainage network for Singapore was obtained from the Public Utilities Board as a key breeding site for mosquitoes around HDBs. The average distance of each HDB block within a sector to a drain was measured as well as the length of the network within the sector^8^.

**Meteorological data** For meteorological data, well-established variables which are known to affect mosquito survival or fecundity were collected. These included daily mean, maximum, and minimum temperature, total rainfall, maximum rainfall falling within a 30-minute, 60-minute and 120-minute window, and wind speed, which were obtained from a total of 21 weather stations installed by the National Environment Agency. We created daily complete raster maps through inverse distance weighting interpolation, which was carried out using cross validation of leave-one-out for the fitting of the inverse distancing power to minimise the error in observation on the raster surface of the test point. Hourly dewpoint and ambient ground air temperature were taken from ERA5, published by ECMWF^9^, to estimate relative humidity over the time period using standard formula. These values were aggregated at a weekly level to correspond with the dengue case data.

**Entomological data** Gravitraps were placed along the common corridors of public housing apartments in all intervention sites, at the ratio of 1 trap for every 20 households, i.e. two to three traps on each of three floors per apartment block: lower floor (2nd), mid floor (5th or 6th floor) and high floor (10th or 11th floor) from EW8 2019 to EW26 2022. This corresponded to an average of six Gravitraps per apartment block. This trap-to-household ratio and deployment was based on logistic considerations for long term monitoring, and it provided an assessment of the density as well as vertical distribution of the mosquito population. Mosquito data was collected from all Gravitraps on a bi-weekly basis.

**Epidemiological data** All individuals who have suspect dengue illness and are tested for dengue, across all public hospitals, general practitioner clinics and polyclinics in Singapore over the study period were eligible to be included. Participants were tested using either using an internally controlled RT-qPCR assay, dengue non-structural protein 1 (NS1) or IgM as diagnostic assays to detect dengue virus in plasma samples from all dengue-suspected patients^10^. Dengue is a notifiable disease in Singapore and incidence data is collected by the Ministry of Health (MOH) for all virologically confirmed cases.

We excluded individuals who were tested on more than one occasion in 4 weeks, individuals who had more than one residential address in different control or intervention townships, individuals who had been tested at different labs with conflicting dengue results. We also excluded individuals who had residential addresses at intervention sites at the time of the test but were not qualified as exposed to *Wolbachia* interventions for at least 3 months based on exposure criteria described below.

**Intervention**

**Spatial characterisation** 117 sectors grouped into 4 intervention towns comprising of an at-risk population of 607,872 residents have been subject to *Wolbachia* releases under two separate approaches. First, the phased release approach where releases were gradually expanded to encompass the sectors in Yishun and Tampines townships (SI Figure 1,2). Secondly, the targeted approach where large areas of the intervention site were subject to *Wolbachia* releases from the start date of intervention. This was employed in Bukit Batok (SI Figure 3) and Choa Chu Kang (SI Figure 4) townships.

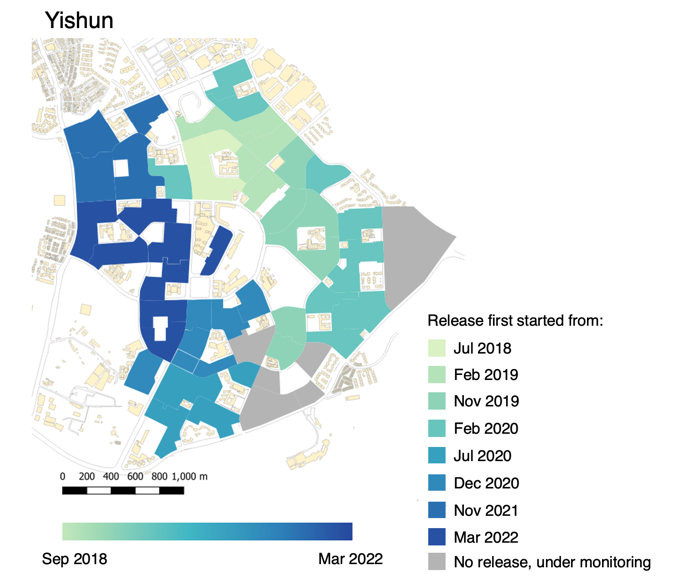

SI Figure 1: Visualization of the phased release approach in Yishun township from Sep 2018 to March
2022. Regions which experienced releases are shaded in the map

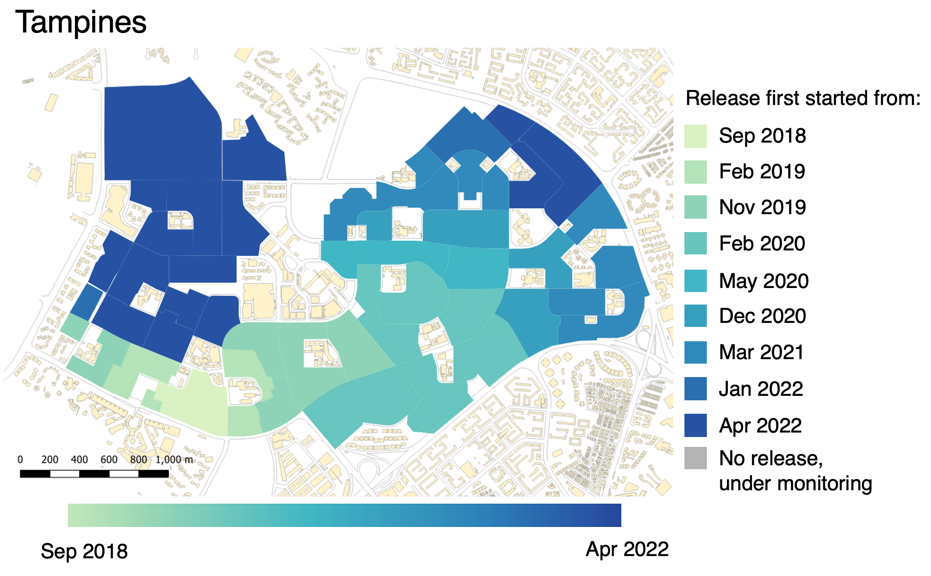

SI Figure 2: Visualization of the phased release approach in Tampines township from Sep 2018 to March
2022. Regions which experienced releases are shaded in the map

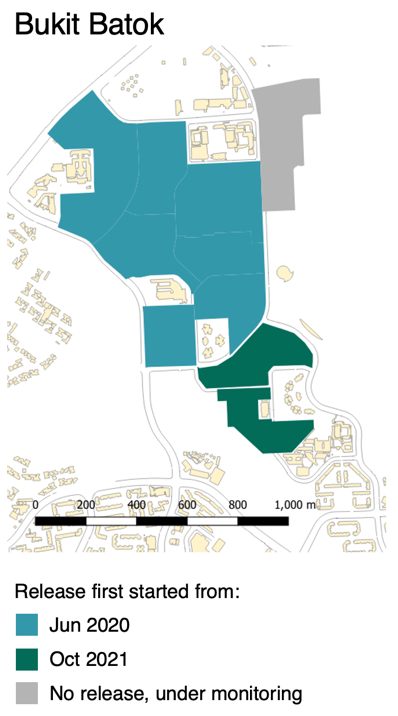

SI Figure 3: Visualization of the targeted release approach in Bukit Batok township from Sep 2018 to March
2022. Regions which experienced releases are shaded in the map

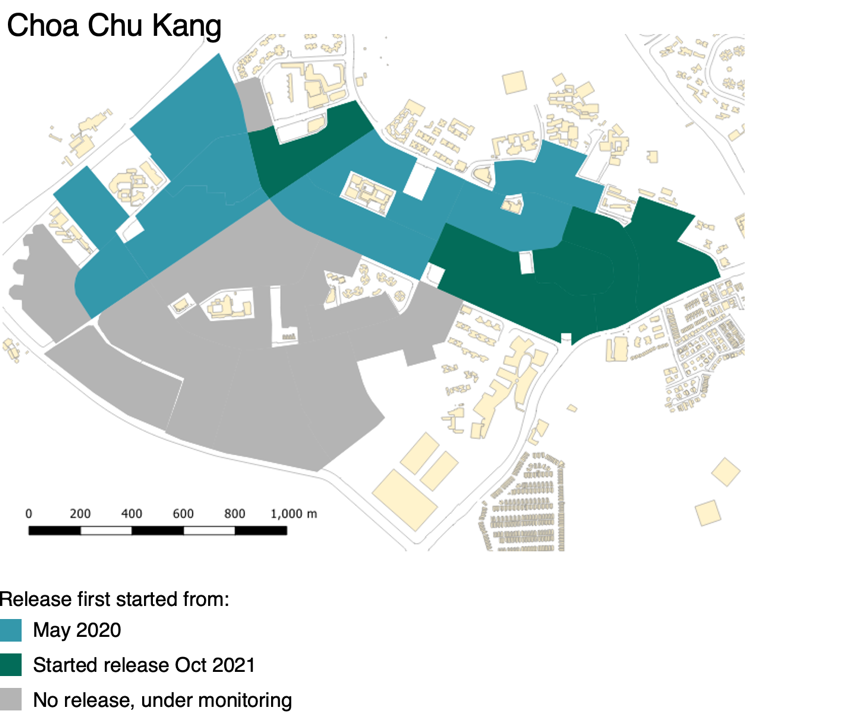

SI Figure 4: Visualization of the targeted release approach in Bukit Batok township from Sep 2018 to March
2022. Regions which experienced releases are shaded in the map

**Monitoring**

Adult *Aedes aegypti* and *albopictus* populations in release and control sectors were monitored weekly using an average of six Gravitraps^11^ per public housing apartment block. Gravitraps were placed in public spaces along corridors and were evenly vertically distributed throughout the block, corresponding to a ratio of approximately one trap for every 20 households. A total of 57,990 traps were deployed in Singapore public housing estates.

All mosquito samples were sexed and morphologically identified, and all *Aedes aegypti* samples were subjected to PCR for *w*AlbB detection in Phase 2 (Before Feb 2019). For Phase 3 onwards after Feb 2019, only female *Aedes aegypti* mosquitoes are subjected to PCR for *w*AlbB detection, whereas female *Aedes* species undergo PCR for molecular identification and *w*AlbB detection. This procedure was continued till the end of the study period.

**Molecular identification of *Aedes* species, wAlB detection and quantification *w*AlbB detection and quantification**

Individual mosquitoes were homogenized in a Beadbeater (Biospec, USA) and total DNA was extracted using the salt lysis extraction method (O’Neill et al., 1992). Female *Aedes* mosquitoes that cannot be speciated morphologically were identified using a multiplex *Ae. aegypti*- and *Ae. albopictus*-specific real-time qPCR assay targeting the *COI* gene (*Aedes* forward 5’-TCC CGC CTT CRG TGC GCG G-3’, *Aedes* reverse 5’-CGC GGG ATG TAY TCA TCA ACC-3’, probe AEG Cy5 – 5’-TAG TCA GAC GTG GTG GTG ACA CAC C-3’– BHQ2, and ALBO HEX – 5’-ACG GTG GCC GGC GTG CCA GTC GT-3’ – BHQ1). Moreover, the detection of *Wolbachia* from field caught female *Aedes aegypti* mosquitoes, targeting the *wsp* gene (*w*AlbB Forward 5’-AGY TAT GAT GTA ACY CCA GA-3’, *w*AlbB Reverse 5’-TTA AAC GCT ACT CCA GCT TCT-3’ and probe *w*ALbB FAM – 5’-T(+G)G TTC TTA T(+G)G TGC TAG TTT TAA TAA A-3’ – BHQ1), can also be incorporated into this multiplex qPCR assay. The multiplex qPCR reactions were performed in 20µl total volume containin 1X SensiFAST No-Rox Probe Mastermix (Bioline, USA); 0.15µM and 0.05µM COI primers and probes, respectively; 0.4 µM and 0.15µM wsp primers and probes, respectively; and 2µl. Cycling was carried out in a Rotorgene Q (Qiagen, Germany) using the following conditions: 95°C for 5 min, 40 cycles at 95°C for 10s, and 60°C for 30 sec.

Quality controls are regularly carried out in our *Ae. aegypti* colonies that are infected with Wolbachia. Wolbachia in our mosquito colonies were quantified using a multiplex real-time qPCR assay targeting the *w*AlbB *wsp* gene (using primers and probe sequences mentioned above) and the *Aedes aegypti* *RpS17* gene (forward primer 5’-TCC GTG GTA TCT CCA TCA AGC T-3’, reverse primer 5’- CAC TTC CGG CAC GTA GTT GTC-3’, probe Texas Red – 5’-CAG GAG GAG GAA CGT GAG CGC AG-3 – BHQ2). qPCR reactions were performed in 20ul total volume containing 1X SensiFAST No-Rox Probe Mastermix (Bioline, USA); 0.4µM and 0.2µM wsp primers and probes, respectively; 0.2µM and 0.1µM RpS17 primers and probes, respectively; and 5µl of DNA template. Cycling was carried out in a LightCycler®96 (Roche, USA) using the following conditions: 95°C for 5 min, 45 cycles at 95°C for 10s, and 60°C for 30 sec. Amplification of the *wsp* and *RpS17* genes from individual mosquitoes was compared against a standard curve generated from a 10-fold serial dilution of DNA standard for each gene.

**Production**
Eggs were hatched as described above. After 24 hours, larvae were counted using an automated larvae counter (Orinno Technology Pte Ltd, Singapore) and transferred to rearing trays measuring (53 x 40.5 x 10 cm) or (103 x 63 x 3 cm) at a density of one or four larvae per ml of deionized water, respectively. Larvae were fed with ground Tetramin Flakes (Tetra, Germany) in either slurry or capsule form. Six days post-hatching, male pupae were separated from female pupae and larvae using the Fay-Morlan glass plate separation method (Wolbaki, China) (Phase 1-2) or the Pupae Separation System (Orinno Technology Pte Ltd, Singapore) prior irradiation for IIT-SIT releases. Pupae were allowed to eclose inside release containers. For releases in Tampines, the automated, multi-step sex separation process described in Crawford et al.^12^ was followed with pupae collected six-days post hatching. In Tampines town SIT protocols were adopted and irradiated mosquitoes were released from Jan 2020 (at a small location). Irradiated mosquitoes were released in the rest of Tampines town from Aug 2020 onwards.

Irradiation was performed with either the RS2000 (Apr 2018 – Jan 2019) or RS2400V irradiators (After Jan 2019) (Rad Source Technologies Inc., USA). For IIT-SIT irradiation (female sterilization), pupae were irradiated in plastic petri dishes with diameter 15.0 cm or in plastic containers measuring 6.5 cm in diameter and 7.0 cm in height (for release). For SIT irradiation (male sterilization), pupae were irradiated in petri dishes with diameter 9.4 cm. Petri dishes or containers were placed at the center of the irradiation chamber of the X-ray irradiator. The effective dose was 30 Gy to 40 Gy, depending on pupae density, irradiator model and desired sterility.

**Emulating constrained randomisation protocol to select control sites**

In a randomized controlled trial, intervention units were pre-selected prior to treatment/placebo allocation, however, long-term field trial townships were not randomly pre-selected for *Wolbachia* interventions. The use of all untreated townships would mean potentially imbalanced historical (and future) dengue incidence rates and transmission risk, thereby making odds of testing positive for dengue skewed between arms. We therefore emulated a constrained randomisation approach for cluster-randomized controlled trials to randomly select 12 control sites from the pool of control townships where there are public housing estates. This prevents potential chance imbalance and selection bias in the generation of a control arm.

**Constrained randomisation protocol** We only include townships which have public housing estates as potential members of the control arm (n=19 of 30), as the 4 field trial sites were public housing towns. In each iteration, we randomly drew 12 control sites as the candidate control arm. The candidate control arm was taken as eligible for analysis if it had similar average historical dengue incidence rate as the 4 control townships from EW1 2010 – EW52 2016. The threshold taken for eligibility was a less than 5% difference in the dengue incidence rates normalized by population size between the intervention and control arms.

1000 random draws were taken, with the balance in historical dengue incidence rates reassessed in each iteration. 411 draws accepted the pool of potential candidate control arms. We randomly selected one constrained randomised selected control arm for main analysis.

As a separate sensitivity analysis, we iteratively generated eligible control arms 1000 times using the constrained randomisation procedure as a robustness check for the stability of our intervention efficacy estimates. This helped ensure that the constrained randomisation protocol produced control arms where intervention efficacy estimates were not affected by the aforementioned procedure (See robustness checks below).

**Statistical Analysis Plan**

**Accounting for confounders and the propensity of *Wolbachia* treatment in both intervention and control arms** We employed a doubly-robust approach to control for any residual differences in covariates which may affect the propensity for each sector to be treated, as well as confound the propensity of an individual to be test positive or negative.

First, we algorithmically pre-selected variables which affected *Wolbachia* exposure status (3,6 and 12 or more months) in the post-intervention period. Prespecified variables included age and gender. Other factors were first selected using the least absolute shrinkage and selection operator (LASSO), with the optimal penalty parameter selected through 5-fold cross validation. LASSO with the optimal penalty parameter was then re-estimated to examine which parameters were not shrunken to 0. We additionally used the variance inflation factor (VIF), with cut off set at 2 to exclude covariates that display high multi-collinearity. The retained parameters were then used for the propensity scoring model as described below.

To control for the propensity of treatment for each individual in a specified sector, we took the dependent variable as whether an individual was *Wolbachia* exposed at 3,6 or 12 or more months respectively, with explanatory variables including the pre-selected age, sex, anthropogenic and environmental variables using the aforementioned variable selection procedure. Thereafter, propensity scores were predicted for each individual using the logistic regression, with inverse probability weights computed as 1/propensity scores for treated, 1/(1-propensity scores) for untreated individuals. We checked for standardized mean differences (SMD) in covariates between groups post-weighting to assess covariate balance. An SMD>0.1 indicates good balance between groups.

**Intention-to-treat analyses: reduction in risk of contracting dengue due to direct *Wolbachia* intervention as estimated by individual level logistic regression**

We took the outcome variable of interest as the dengue test positive/negative status per individual. The exposure variable of interest was *Wolbachia* release at an individual’s place of residence (home address) point of testing. Here, we reclassified *Wolbachia* exposure as 3, 6 or 12 or more months of *Wolbachia* exposure. This is based off prior data which showed that 6 or more months is required to achieve 80% suppression in *Ae aegypti* abundance. We ran a doubly logistic regression, while controlling for same variables used to estimate propensity scores in first step, to control for any potential model misspecification in either step.

Thereafter, the regression coefficient for *Wolbachia* exposure can be obtained, and its accompanying odds ratio (OR) can be used to compute the protective efficacy of the intervention. An OR<1 denotes that *Wolbachia* exposure has a protective effect on an individual’s risk of contracting dengue. We defined protective efficacy as (1-OR) x 100. Inference was conducted using the cluster bootstrap approach^13^, where we randomly resampled our dataset based on sector or township to construct the empirical distribution of regression coefficients. Correspondingly, we used the percentile method to obtain 95% confidence intervals. We deem the OR significant if the 95% CI is not bounded by 1.

**Intention-to-treat analyses: protective efficacies by subgroup** We re-ran our intention-to-treat analysis by subgroup. Namely, we subsetted to age groups to paediatrics/adolescents (0–18), adults (18–65) and elderly (65+), by male and female sexes and reran the logistic regression procedure as detailed above. We also reran our analysis based on year (2019, 2020, 2021, 2022) to look at the impact of the intervention on interepidemic/epidemic years, and township (Bukit Batok, Choa Chu Kang, Tampines, Yishun) to examine site-specific protective efficacies.

**Robustness Checks**

**Results for robustness checks (1 of 5) on constrained randomisation scheme** Our main estimates relied only on one random set of control townships as the control arm and it may be due to chance that estimated intervention effects were due to chance and control selection. We reran our intention-to-treat analysis on the main cohort, iteratively taking the 1000 eligible potential control arm sets as the control group to examine the robustness of our main estimates under different constrained allocations.

We showed that there was no major deviation in the aggregate protective efficacies versus the pre-selected control arm (SI Figure 5, SI Table 2). All protective efficacies were positive and significant at the 95% level, with the grand mean being slightly higher compared to our point estimates – implying that our point estimates of protective efficacy are conservative, relative to the set of estimated protective efficacies under different constrained randomisations.

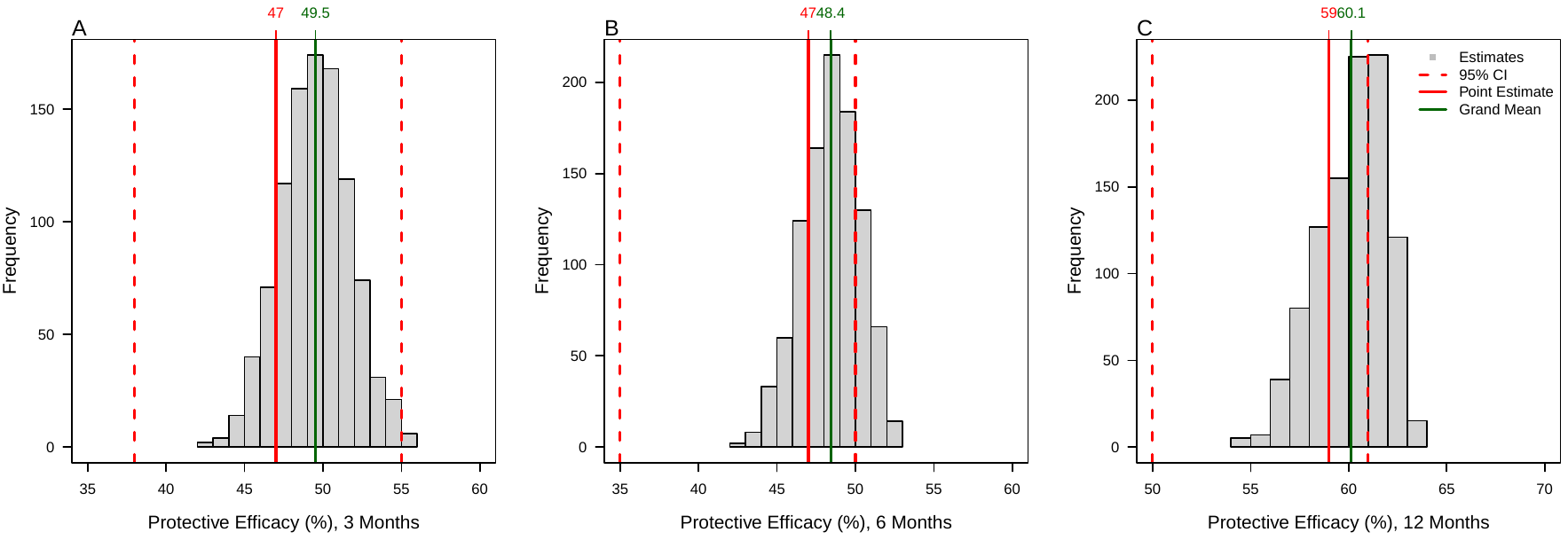

**SI Figure 5:** Histogram of protective efficacies for 1000 eligible potential control arms compared against the point estimate of the randomly selected control allocation and the accompanying 95% confidence intervals of that point estimate. Protective efficacies for Wolbachia exposure at (**A**) 3 or more months, (**B**) 6 or more months and (**C**) 12 or more months were visualized.

|  | Wolbachia Exposure (Months) | | |
| --- | --- | --- | --- |
| Summaries | 3+ | 6+ | 12+ |
| OR (Minimum) | 0.443 | 0.471 | 0.364 |
| OR (Median) | 0.504 | 0.515 | 0.396 |
| OR (Mean) | 0.505 | 0.516 | 0.399 |
| OR (Maximum) | 0.577 | 0.579 | 0.458 |
| OR Point Estimate (95% CI) | 0.53 [0.45-0.62] | 0.53 [0.50-0.65] | 0.41 [0.39-0.50] |

**SI Table 2**: Summary statistics for Odds Ratios (ORs) for 1000 eligible potential control arms compared against the point estimate of the randomly selected control allocation and the accompanying 95% confidence intervals of that point estimate. All estimates relied on the doubly-robust logistic regression with algorithmically selected covariates.

**Results for robustness checks (2 of 5) on covariate selection and inverse probability weighted analyses without adjustment for covariates in the main logistic regression step** We repeated all analyses for the doubly-robust logistic regression taking the full set of covariates as explanatory variables instead of the algorithmically selected covariates, to recompute ORs for protective efficacy. We found no large differences in estimated ORs with or without incorporation of the full covariate set. We also repeated all analysis without incorporation of covariates in the main logistic regression step to examine the influence of using the doubly-robust logistic regression versus a normal inverse-probability weighted analysis. We found no large differences in estimated ORs between both approaches (SI Table 3).

|  | Unadjusted^#^ | Adjusted for all covariates^##^ | Exposed^###^ | Unexposed^####^ |
| --- | --- | --- | --- | --- |
| Exposure Time | OR  (95% CI) | | Test-positive (%)^**^ | |
| By year | Aggregate | | | |
| 3 months + | 0.57^*^ (0.52-0.71) | 0.54^*^ (0.51-0.63) | 13.6% (956/7049) | 21.7%  (14986/69216) |
| 6 months + | 0.56^*^  (0.52-0.68) | 0.54^*^  (0.51-0.65) | 12.9% (767/5935) | 21.6%  (15175/70330) |
| 12 months + | 0.40^*^ (0.37-0.52) | 0.42^*^  (0.39-0.50) | 10.5% (428/4077) | 21.5% (15514/72188) |

SI Table 3: Odds ratios (OR) and test positive percentages for different *Wolbachia* exposure categories and across year, town, age and sex subgroups.

^*^Denotes an OR < 1 with 95% confidence intervals (CI) which are not bounded by 1 denotes a significant protective effect of *Wolbachia* interventions on the risk of dengue.

^#^ORs were estimated using logistic regression with weights for each individual estimated using inverse probability weighting. Cluster bootstrap at the sector resolution was used to obtain CIs to account for sector-specific spatial clustering of data and the intervention.

^##^ORs were estimated using doubly logistic regression with weights for each individual estimated using inverse probability weighting adjusted for all covariates available. Cluster bootstrap at the sector resolution was used to obtain CIs to account for sector-specific spatial clustering of data and the intervention.

^###^An individual testing for febrile illness is considered *Wolbachia*-exposed if the individual resides in a sector with 3,6 or 12 or more months of sustained *Wolbachia* release

^####^Unexposed group taken as the pre-selected, constrain randomised set of 12 controls

^**^Unweighted percentages of individuals testing positive in *Wolbachia*-exposed and *Wolbachia*-unexposed sectors

**Results for robustness checks (3 of 5) using in-space placebo checks on control sites** We iteratively took half of the allocated control sites as the allocated placebo-intervention site and re-estimated odds ratios and intervention efficacies by comparing test-negative and positive individuals in the placebo-intervention versus other the remaining control sites 100 times. If our estimated protective efficacy was only demonstrated in the true intervention sites, then there should be no significant differences between the pseudo-intervention and control sites. This was done to better ensure that we did not obtain spurious intervention efficacy estimates using our estimation procedure. Placebo mean ORs were far smaller compared to the actual intervention efficacy using the in-space placebo check (SI Figure 6, SI Table 4).

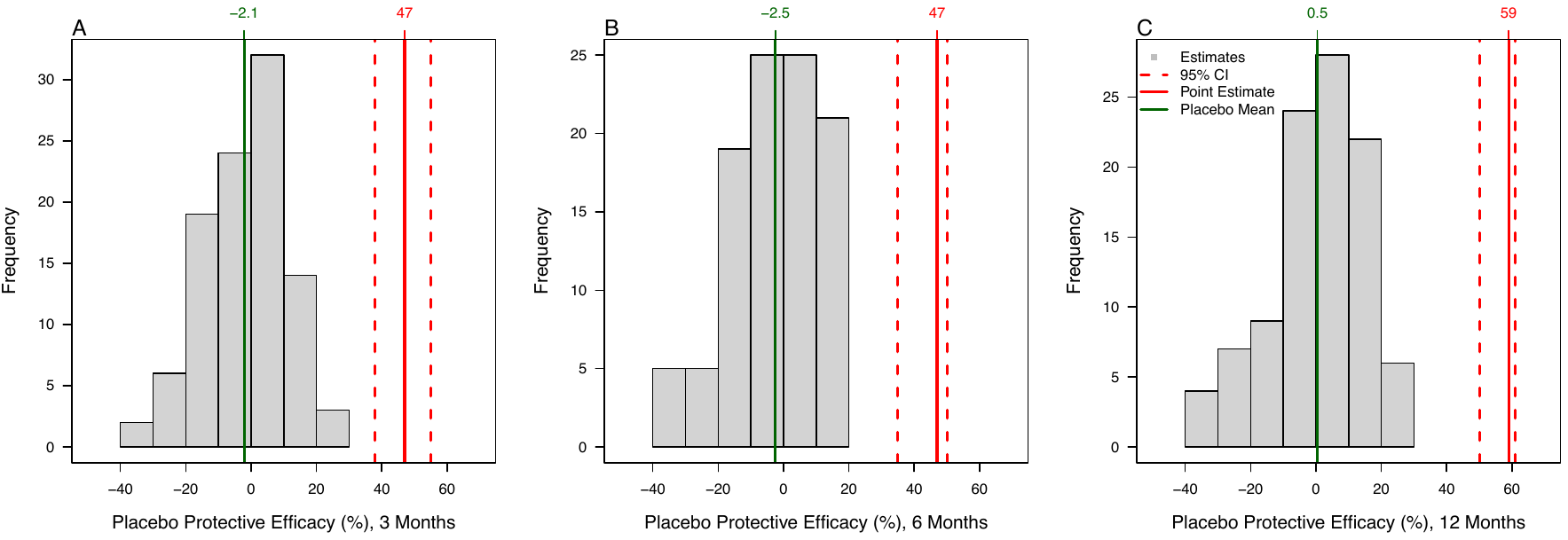

**SI Figure 6:** Histogram of placebo protective efficacies for 100 random placebo-intervention and control arms compared against the point estimate of the randomly selected control allocation and the accompanying 95% confidence intervals of that point estimate. Protective efficacies for Wolbachia exposure at (**A**) 3 or more months, (**B**) 6 or more months and (**C**) 12 or more months were visualized.

| Exposure | Significant ORs > 1 | Significant ORs < 1 | Placebo OR Grand Mean | Placebo OR Range | Proportion ORs >  Point Estimate OR |
| --- | --- | --- | --- | --- | --- |
| 3 months | 42 | 16 | 1.021 | 0.718, 1.392 | 0 |
| 6 months | 42 | 20 | 1.025 | 0.802, 1.379 | 0 |
| 12 months | 34 | 20 | 0.995 | 0.710, 1.340 | 0 |

**SI Table 4**: Summary statistics for Odds Ratios (ORs) for 100 random placebo-intervention and control arms compared against the point estimate of the randomly selected control allocation and the accompanying 95% confidence intervals of that point estimate.. All estimates relied on the doubly-robust logistic regression with algorithmically selected covariates.

**Results for robustness checks (4 of 5) using in-time placebo checks on control sites** We conducted placebo checks on intervention sites, taking each intervention site in the periods before the actual intervention as the intervention period, and recomputed intervention efficacies among allocated controls and interventions based on this placebo-intervention period. If our estimated protective efficacy was only demonstrated in the true intervention sites in the post-intervention period, then there should be no significant differences between intervention and control sites in the pre-intervention period. This was done to better ensure that we did not obtain spurious intervention efficacy estimates using our estimation procedure. We found no significant ORs in the in-time exposure for intervention sites in the pre-intervention period using the in-time placebo check.

|  | Placebo Intervention Period | | | | | |
| --- | --- | --- | --- | --- | --- | --- |
|  | EW 1 2016 – EW 27 2018 | | | EW 1 2016 – EW 52 2018 | | |
| Placebo *Wolbachia*  Exposure | OR | 95% CI LB | 95% CI UB | OR | 95% CI LB | 95% CI UB |
| 3+ | 1.044 | 0.986 | 1.287 | 1.041 | 0.985 | 1.278 |
| 6+ | 1.030 | 0.972 | 1.276 | 1.028 | 0.965 | 1.280 |
| 12+ | 1.008 | 0.945 | 1.241 | 1.017 | 0.957 | 1.243 |

**SI Table 5**: Summary statistics for Odds Ratios (ORs) for in-time placebo check for actual intervention and control arms and the accompanying 95% confidence intervals of that point estimate.. All estimates relied on the doubly-robust logistic regression with algorithmically selected covariates.

**Results for robustness checks (5 of 5) Results: cluster-bootstrap inference using towns instead of sectors** We obtained point estimates and confidence intervals using the cluster-bootstrap approach but with town-level clustering instead of sector level clustering. 95% CIs were wider using town-level clustering assumption, no large differences in significance were found between this and the sector-level assumption (SI Table 6).

|  | OR  (95% CI)* | Test-positive  (%)^##^ | | OR  (95% CI) | Test-positive  (%) | | OR  (95% CI) | Test-positive  (%) | | OR  (95% CI) | Test-positive  (%) | | OR  (95% CI) | Test-positive  (%) | |
| --- | --- | --- | --- | --- | --- | --- | --- | --- | --- | --- | --- | --- | --- | --- | --- |
| Exposure Time^#^ |  | Exposed | Unexposed |  | Exposed | Unexposed |  | Exposed | Unexposed |  | Exposed | Unexposed |  | Exposed | Unexposed |
| By year | **2019** | | | **2020** | | | **2021** | | | **2022** | | | **Aggregate** | | |
| 3 months + | **0.52*  (0.42-0.74)** | 7.7%  (37/482) | 17.2% (3839/22377) | **0.59*  (0.27-0.88)** | 21.9% (525/2395) | 27.5% (6549/23852) | 0.73  (0.27-2.14) | 6.4%  (147/2294) | 9.5% (1121/11787) | **0.37*  (0.31-0.57)** | 13.2% (247/1878) | 31.0% (3477/11200) | **0.53*  (0.30-0.78)** | 13.6% (956/7049) | 21.7%  (14986/69216) |
| 6 months + | **0.38* (0.29-0.56)** | 6.1%  (19/314) | 17.1% (3857/22545) | **0.64*  (0.53-0.86)** | 22.4%  (376/1679) | 27.3% (6698/24568) | 0.68  (0.60-2.39) | 6.3%  (137/2191) | 9.5% (1131/11890) | **0.41*  (0.35-0.66)** | 13.4% (235/1751) | 30.8% (3489/11327) | **0.53*  (0.44-0.75)** | 12.9% (767/5935) | 21.6%  (15175/70330) |
| 12 months + | **0.22*  (0.09-0.35)** | 3.2%  (3/94) | 17.0% (3873/22765) | **0.50*  (0.45-0.61)** | 15.9%  (98/617) | 27.2%  (6976/25630) | **0.52*  (0.44-0.92)** | 6.2% (108/1731) | 9.4% (1160/12350) | **0.40*  (0.35-0.66)** | 13.4% (219/1635) | 30.6% (3505/11443) | **0.41*  (0.36-0.66)** | 10.5%  (428/4077) | 21.5%  (15514/72188) |
| By Township | **Bukit Batok** | | | **Choa Chu Kang** | | | **Yishun** | | | **Tampines** | | |  |  |  |
| 3 months + | 0.46  (0.44-1.05) | 11.0%  (42/382) | -^**^ | 0.65  (0.63-1.48) | 16.2% (137/845) | -^**^ | **0.32*  (0.30-0.52)** | 9.1% (286/3127) | -^**^ | **0.62*  (0.53-0.74)** | 18.2% (491/2695) | -^**^ |  |  |  |
| 6 months + | 0.43  (0.44-1.08) | 11.4% (37/325) | -^**^ | 0.68  (0.67-1.27) | 15.4% (105/682) | -^**^ | **0.31*  (0.30-0.47)** | 8.2% (219/2676) | -^**^ | **0.63*  (0.56-0.73)** | 18.0% (406/2252) | -^**^ |  |  |  |
| 12 months + | **0.45*  (0.41-0.67)** | 13.4% (29/216) | -^**^ | **0.64*  (0.58-0.75)** | 13.3% (59/443) | -^**^ | **0.23*  (0.22-0.29)** | 8.0% (155/1937) | -^**^ | **0.49*  (0.46-0.56)** | 12.5%  (185/1481) | -^**^ |  |  |  |
| By age and gender | **0-20** | | | **20-65** | | | **65+** | | | **Male** | | | **Female** | | |
| 3 months + | **0.57*  (0.39-0.91)** | 12.4% (93/750) | 18.4% (1881/10249) | **0.54*  (0.41-0.76)** | 16.5% (664/4033) | 24.8% (10427/42039) | 0.53  (0.47-1.00) | 8.7% (195/2237) | 15.5% (2584/16637) | **0.67*  (0.47-0.97)** | 14.6% (497/3397) | 22.2% (7098/32017) | **0.39*  (0.35-0.52)** | 11.1%  (373/3371) | 19.6% (6008/30648) |
| 6 months + | 0.58  (0.41-1.03) | 12.7% (80/630) | 18.3% (1894/10369) | **0.53*  (0.46-0.68)** | 18.5% (525/2842) | 22.7% (10566/42705) | 0.55  (0.47-1.84) | 8.3% (158/1912) | 15.5% (2621/16962) | 0.62  (0.48-1.02) | 13.7% (392/2861) | 22.1% (7203/32551) | **0.44*  (0.39-0.61)** | 10.8%  (308/2842) | 19.5% (6073/31177) |
| 12 months + | **0.38*  (0.26-0.71)** | 8.9% (38/425) | 18.3% (1936/10574) | **0.40*  (0.35-0.55)** | 12.5%  (283/2271) | 24.7% (10808/43801) | 0.51  (0.43-1.71) | 7.7% (105/1358) | 15.3% (2674/17516) | **0.50*  (0.44-0.81)** | 11.9% (238/1999) | 22.0 (7357/33415) | **0.29*  (0.26-0.42)** | 8.3% (161/1943) | 19.4% (6220/32076) |

SI Table 6: Odds ratios (OR) and test positive percentages for different *Wolbachia* exposure categories and across year, town, age and sex subgroups.

^*^Denotes an OR < 1 with 95% confidence intervals (CI) which are not bounded by 1 denotes a significant protective effect of *Wolbachia* interventions on the risk of dengue. ORs are estimated using doubly robust logistic regression with weights for each individual estimated using inverse probability weighting. Cluster bootstrap at the town resolution was used to obtain CIs to account for sector-specific spatial clustering of data and the intervention.

^**^Unexposed group taken as the pre-randomised set of 12 controls

^#^An individual testing for febrile illness is considered *Wolbachia*-exposed if the individual resides in a sector with 3,6 or 12 or more months of sustained *Wolbachia* release

^##^Unweighted percentages of individuals testing positive in *Wolbachia*-exposed and *Wolbachia*-unexposed sectors

**Summary statistics for clustered versus sporadic annual dengue incidence rates** Clustered cases were taken as those which reside in a dengue cluster–defined as two or more cases with onset days within 14 days, located within 150 m of each other based on their house address, according to the operational criteria of the National Environment Agency, Singapore. Sporadic cases are defined as those who do not reside in a dengue cluster. We normalized incidence at the town level by the number of Singapore residents in the release area of each town multiplied by 100,000 to obtain incidence per 100,000 persons (incidence rate) from EW1 2014–EW26 2022. This is summarized in SI Table 7 below.

|  | Pre-intervention  (EW1 2014 - EW52 2018) | | Post-intervention   (EW1 2019 - EW26 2022) | |
| --- | --- | --- | --- | --- |
|  | Mean annual dengue incidence rate | SD | Mean annual dengue incidence rate | SD |
|  | **Sporadic** | | | |
| *Study Towns* |  |  |  |  |
| Bukit Batok | 49.84 | 23.70 | 41.11 | 18.92 |
| Choa Chu Kang | 56.59 | 26.80 | 50.25 | 17.61 |
| Tampines | 50.80 | 28.56 | 40.71 | 8.27 |
| Yishun | 46.75 | 23.34 | 39.61 | 14.53 |
| Intervention | 50.99 | 23.86 | 42.92 | 14.45 |
| *Control Towns* |  |  |  |  |
| Bedok | 84.78 | 50.94 | 63.09 | 24.16 |
| Bishan | 64.20 | 37.75 | 64.58 | 29.38 |
| Clementi | 51.52 | 27.91 | 50.66 | 28.07 |
| Geylang | 112.00 | 79.33 | 61.46 | 20.77 |
| Jurong West | 55.44 | 30.48 | 44.97 | 17.85 |
| Kallang | 57.10 | 30.35 | 45.45 | 10.87 |
| Pasir Ris | 69.29 | 40.99 | 59.97 | 22.01 |
| Punggol | 31.23 | 12.49 | 36.34 | 13.55 |
| Queenstown | 46.98 | 23.89 | 37.48 | 15.78 |
| Sengkang | 44.87 | 23.17 | 39.62 | 13.89 |
| Toa Payoh | 54.91 | 33.91 | 45.19 | 12.74 |
| Woodlands | 58.58 | 27.56 | 37.41 | 18.41 |
| Control | 60.91 | 40.13 | 48.85 | 20.24 |
|  | **Clustered** | | | |
| *Study Towns* |  |  |  |  |
| Bukit Batok | 20.93 | 23.55 | 105.90 | 108.59 |
| Choa Chu Kang | 55.97 | 89.95 | 158.49 | 138.74 |
| Tampines | 85.06 | 64.40 | 118.09 | 132.56 |
| Yishun | 76.27 | 64.30 | 71.75 | 65.28 |
| Intervention | 59.56 | 64.84 | 113.56 | 107.70 |
| *Control Towns* |  |  |  |  |
| Bedok | 164.07 | 114.89 | 355.02 | 312.37 |
| Bishan | 161.03 | 241.40 | 313.96 | 271.84 |
| Clementi | 31.08 | 26.21 | 147.47 | 182.28 |
| Geylang | 249.10 | 256.10 | 566.05 | 613.29 |
| Jurong West | 55.11 | 31.33 | 201.52 | 142.32 |
| Kallang | 98.87 | 92.84 | 198.79 | 209.02 |
| Pasir Ris | 123.22 | 96.77 | 237.51 | 206.04 |
| Punggol | 10.88 | 6.35 | 42.32 | 37.35 |
| Queenstown | 13.82 | 6.57 | 200.02 | 160.90 |
| Sengkang | 30.31 | 31.43 | 118.31 | 85.73 |
| Toa Payoh | 63.27 | 59.89 | 279.77 | 269.85 |
| Woodlands | 48.23 | 28.30 | 286.41 | 201.02 |
| Control | 87.42 | 126.71 | 245.60 | 264.48 |

SI Table 7: summary statistics for mean annual dengue incidence rate and standard deviations (SD) by town and by NEA operational case definitions (clustered/sporadic).
